# First-line arteriovenous access and risks of hospitalization and death in patients starting hemodialysis – a nationwide cohort study

**DOI:** 10.1101/2022.12.28.22283990

**Authors:** Natalia Alencar de Pinho, Mathilde Prezelin-Reydit, Jerome Harambat, Cécile Couchoud, Florence Glaudet, Christian Combe, Virginie Rondeau, Karen Leffondré, the REIN registry

## Abstract

Arteriovenous (AV) access choice has sparked controversy with recent evidence suggesting overestimation of benefits associated with AV fistula versus graft in certain populations. We assessed outcomes associated with first-line AV access type in patients who started hemodialysis with a catheter in France, overall and by subgroups of age, sex, and comorbidities. In this retrospective cohort study, we included incident patients who initiated hemodialysis with a catheter from 2010 through 2018, followed by the French REIN Registry. Our main exposure was the first-line (first-created) AV graft versus fistula, ascertained through the linkage with the French national health-administrative database. Outcomes were all-cause and cause-specific hospitalization, and all-cause mortality. We used joint frailty models to deal with recurrent hospitalizations and informative censoring by death, Cox proportional hazard (PH) models, and inverse probability weighting. From the 18,625 patients included (mean age was 68±15 years, 35% were women), 5% had a first-line AV graft. Patients with AV graft had an 11%-higher weighted hazard of all-cause hospitalization (95% CI 1.09 to 1.13), 16% higher weighted hazard of cardiovascular (95% CI 1.05 to 1.29) and infection-related (95% CI 1.01 to 1.33) hospitalization, 34% higher weighted hazard of vascular access-related hospitalization, and a 9%-higher weighted hazard of all-cause death (95% CI 0.97 to 1.23). Results were consistent for most subgroups, except that the highest hazard of hospitalization with AV graft was blunted in patients with comorbidities (i.e. diabetes, weighted HR of all-cause hospitalization 1.03, 95% CI 0.95-1.12).- To conclude, in patients starting hemodialysis with a catheter, first-line AV graft is associated with increased hazard of hospitalization vs. patients with AV fistula. This may, however, not be the case for patients with a poor vascular condition, i.e., those with diabetes, who have a similar hospitalization and mortality rates with either graft or fistula.

## Introduction

The arteriovenous access created by Quinton, Dillard, and Scribner^1^ in the early 60’s marked a watershed in the prognosis of patients with irreversible kidney failure, making long-term hemodialysis possible. Further improvements in hemodialysis vascular access, notably with the development of arteriovenous (AV) fistula, AV graft, and central venous catheter, contributed to increasing eligible population for hemodialysis.^2,3^ Today, more than 2 million people are treated with hemodialysis worldwide.^4,5^ Diabetes is a major cause of kidney replacement therapy (KRT) and, in developed countries, both incidence and prevalence of KRT are the highest among the elderly.^6^

In the absence of randomized clinical trials comparing different vascular access types, substantial uncertainty exists regarding the impact of vascular access strategy on patient outcomes. First-line AV fistula have been advocated because of its association with fewer access complications and better patient outcomes in observational studies.^7–9^ However, high rates of maturation failure^10,11^ and longer catheter dependency^12–15^ have been described in patients who underwent AV fistula versus AV graft creation and required a catheter for hemodialysis initiation. This is especially the case of the elderly and patients with high comorbidity burden.

Hesitation to anticipate AV access due to the competing risk of death before KRT start may lead to catheter use at hemodialysis initiation, which has itself been associated with subsequent AV fistula failure.^16–18^ As a consequence, the benefits of AV fistula in terms of patient survival and morbidity have been considered overestimated when assessed through the first access used, as opposed to the first access created.^19^ In this respect, some studies have found comparable patient survival^20^ and hospitalization rates^21^ with either AV fistula or graft in elderly patients starting hemodialysis with a catheter, but not others.^22,23^ Discrepancies between results from previous studies may be explained by differences in the populations included and in the methods used.

In this study, we sought to assess hospitalization and mortality risks associated with first-line AV access type, i.e. the first AV access created, in patients who started hemodialysis in France with a catheter and may thus be at high risk of AV fistula maturation failure, overall and by subgroups of age, sex, and comorbidities.

## Methods

### Study Sample & Data Collection

The French REIN registry includes all patients receiving kidney replacement therapy (KRT) for kidney failure in France. Details on methods and quality control have been described elsewhere.^24^ The REIN registry and its utilization for research purposes have been approved by the relevant French ethics committees, specifically, the Comité consultatif sur le traitement de l’information en matière de recherche (CCTIRS) and the Commission nationale de l’informatique et des libertés (CNIL N° 903188). French regulations do not require participants’ written or verbal informed consent for their inclusion in population-based registries requiring exhaustiveness. Patients are informed about the registration in the REIN registry and their right to not participate (opt out) by the nephrology clinic.

The current study sample includes adult patients (age ≥ 18 years) who initiated KRT with hemodialysis from 2010 through 2018, and who had a first AV access placement (fistula or graft) after hemodialysis initiation. Subgroups of interest were defined by age (<70, 70-79, and ≥80 years), sex, and dichotomous (yes-no) history of diabetes, heart failure and peripheral artery disease. Demographics, primary kidney disease, comorbidities, and other study covariates were recorded by nephrologists or clinical research associates at KRT initiation, at any significant changes in KRT modality, and yearly over registry follow-up. A linkage of the REIN registry with the French health administrative database (SNDS), which assembles information on all reimbursed in- and out of-hospital care in 99% of the population, has been recently implemented.^25^ This allowed us to obtain information on vascular access procedures and hospitalizations occurring two years before and up to 4 years after hemodialysis initiation.

### Exposure

The study exposure was the type of first AV access placed following hemodialysis initiation. AV fistula placement was identified in the SNDS database through two codes (EZMA003 or EZMA001 for *AV fistula creation for vascular access through open surgery with or without superficialization, respectively*) and AV graft placement through one code (EZCA003 for *AV graft creation for vascular access through open surgery*). The absence of previous attempts of AV access placement was assessed through the history of AV access procedures over the two-year period preceding hemodialysis initiation, and by the covariate “date of the first AV fistula creation” in the REIN registry.

### Outcomes

Study outcomes were time from first AV access placement after hemodialysis initiation with a catheter to all-cause mortality, all-cause hospitalization and cause-specific hospitalization. Cause-specific hospitalizations included those related to cardiovascular disease, infectious syndromes, or to vascular access complications, based on hospitalization main ICD-10 code or on the French diagnosis related groups (grouping hospital care for reimbursement purposes, Supplementary Box). Only hospitalizations with at least one overnight stay were considered in the analysis. Patients were followed-up for 48 months or until kidney transplantation, switch to peritoneal dialysis, loss to follow-up, death, or December 31, 2019, whichever occurred first.

### Statistical Analysis

#### Propensities scores of first-line AV fistula or graft

To account for confounding that may arise from non-randomized allocation to AV access groups, we used a propensity score approach in which each patient is weighted by the inverse of one’s probability to receive a first-line fistula or graft.^26^ This probability was estimated with a logistic regression including potential confounders of the relationship between the type of AV access and outcomes (variables in Table 1, according to the framework illustrated by the direct acyclic graph in Figure S1). Further details about the propensity score model is given is Supplementary Methods. Because 36% of patients had at least one missing value on selected confounders, we performed multiple imputation. We generated 36 datasets,^27^ with Proc MI (SAS version 9.4, SAS Institute Inc., Cary, NC) using the full conditional specification method.

**Table 1.**
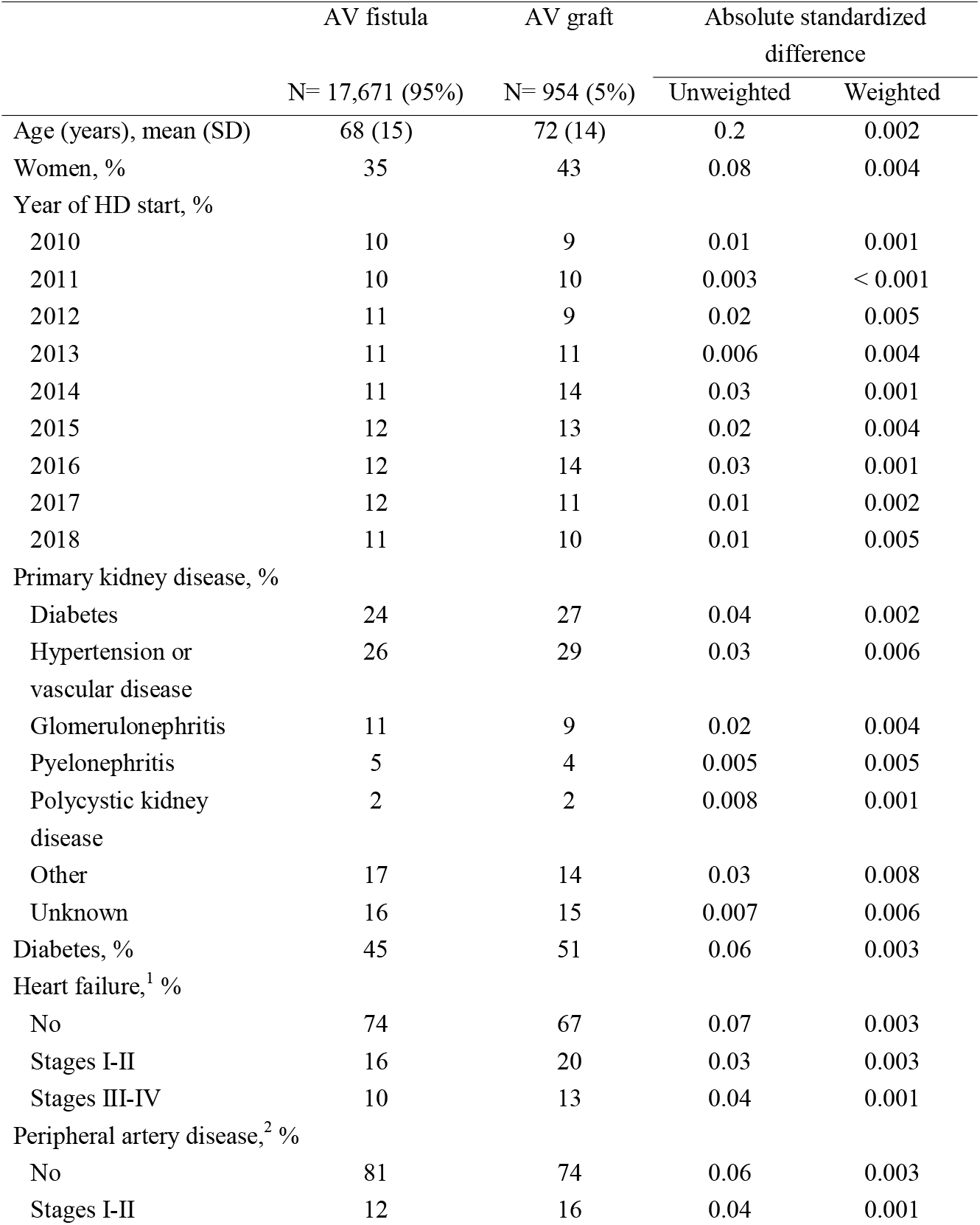
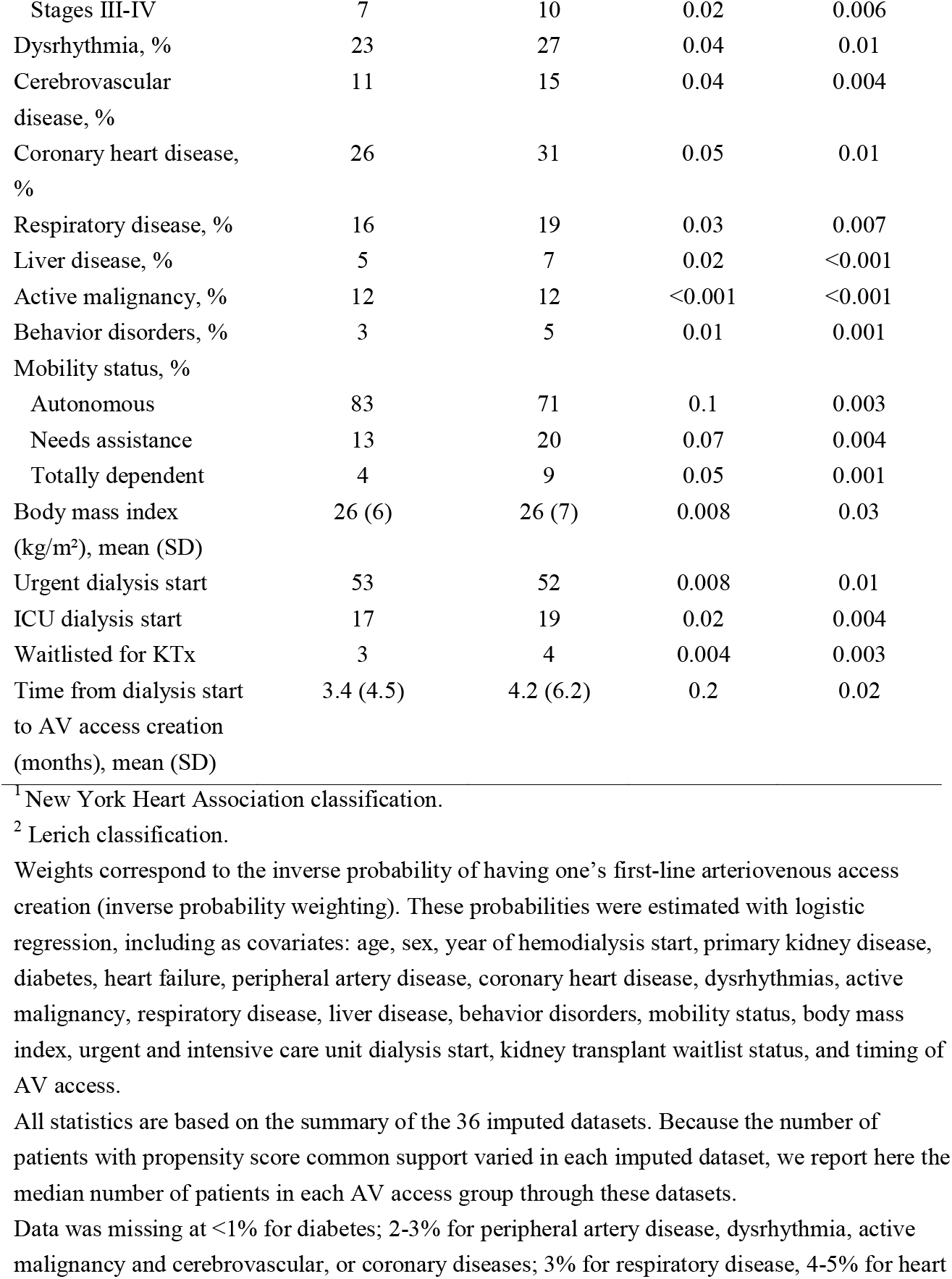
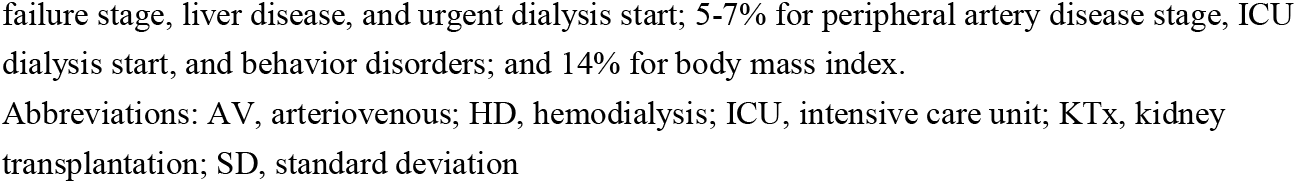
Characteristics of patients by type of first-line AV access.

|  | AV fistula<br>N= 17,671 (95%) | AV graft<br>N= 954 (5%) | Absolute standardized<br>difference |  |
| --- | --- | --- | --- | --- |
|  |  |  | Unweighted | Weighted |
| Age (years), mean (SD) | 68 (15) | 72 (14) | 0.2 | 0.002 |
| Women, % | 35 | 43 | 0.08 | 0.004 |
| Year of HD start, % |  |  |  |  |
| 2010 | 10 | 9 | 0.01 | 0.001 |
| 2011 | 10 | 10 | 0.003 | < 0.001 |
| 2012 | 11 | 9 | 0.02 | 0.005 |
| 2013 | 11 | 11 | 0.006 | 0.004 |
| 2014 | 11 | 14 | 0.03 | 0.001 |
| 2015 | 12 | 13 | 0.02 | 0.004 |
| 2016 | 12 | 14 | 0.03 | 0.001 |
| 2017 | 12 | 11 | 0.01 | 0.002 |
| 2018 | 11 | 10 | 0.01 | 0.005 |
| Primary kidney disease, % |  |  |  |  |
| Diabetes | 24 | 27 | 0.04 | 0.002 |
| Hypertension or<br>vascular disease | 26 | 29 | 0.03 | 0.006 |
| Glomerulonephritis | 11 | 9 | 0.02 | 0.004 |
| Pyelonephritis | 5 | 4 | 0.005 | 0.005 |
| Polycystic kidney<br>disease | 2 | 2 | 0.008 | 0.001 |
| Other | 17 | 14 | 0.03 | 0.008 |
| Unknown | 16 | 15 | 0.007 | 0.006 |
| Diabetes, % | 45 | 51 | 0.06 | 0.003 |
| Heart failure,<sup>1</sup> % |  |  |  |  |
| No | 74 | 67 | 0.07 | 0.003 |
| Stages I-II | 16 | 20 | 0.03 | 0.003 |
| Stages III-IV | 10 | 13 | 0.04 | 0.001 |
| Peripheral artery disease,<sup>2</sup> % |  |  |  |  |
| No | 81 | 74 | 0.06 | 0.003 |
| Stages I-II | 12 | 16 | 0.04 | 0.001 |
| Stages III-IV | 7 | 10 | 0.02 | 0.006 |
| Dysrhythmia, % | 23 | 27 | 0.04 | 0.01 |
| Cerebrovascular disease, % | 11 | 15 | 0.04 | 0.004 |
| Coronary heart disease, % | 26 | 31 | 0.05 | 0.01 |
| Respiratory disease, % | 16 | 19 | 0.03 | 0.007 |
| Liver disease, % | 5 | 7 | 0.02 | <0.001 |
| Active malignancy, % | 12 | 12 | <0.001 | <0.001 |
| Behavior disorders, % | 3 | 5 | 0.01 | 0.001 |
| Mobility status, % |  |  |  |  |
| Autonomous | 83 | 71 | 0.1 | 0.003 |
| Needs assistance | 13 | 20 | 0.07 | 0.004 |
| Totally dependent | 4 | 9 | 0.05 | 0.001 |
| Body mass index (kg/m<sup>2</sup>), mean (SD) | 26 (6) | 26 (7) | 0.008 | 0.03 |
| Urgent dialysis start | 53 | 52 | 0.008 | 0.01 |
| ICU dialysis start | 17 | 19 | 0.02 | 0.004 |
| Waitlisted for KTx | 3 | 4 | 0.004 | 0.003 |
| Time from dialysis start to AV access creation (months), mean (SD) | 3.4 (4.5) | 4.2 (6.2) | 0.2 | 0.02 |

<sup>1</sup> New York Heart Association classification.
<sup>2</sup> Lerich classification.
All statistics are based on the summary of the 36 imputed datasets. Because the number of patients with propensity score common support varied in each imputed dataset, we report here the median number of patients in each AV access group through these datasets.
Data was missing at <1% for diabetes; 2-3% for peripheral artery disease, dysrhythmia, active malignancy and cerebrovascular, or coronary diseases; 3% for respiratory disease, 4-5% for heart
failure stage, liver disease, and urgent dialysis start; 5-7% for peripheral artery disease stage, ICU dialysis start, and behavior disorders; and 14% for body mass index.
Abbreviations: AV, arteriovenous; HD, hemodialysis; ICU, intensive care unit; KTx, kidney transplantation; SD, standard deviation

#### Risks of hospitalization and death associated with first-line AV access

To estimate the adjusted association between AV access and hospitalizations, we used joint weighted semiparametric frailty models. Each joint model included a submodel for the hazard of recurrent hospitalizations (all cause or cause-specific hospitalizations) and a submodel for the hazard of all cause death, which was considered as informative censoring.^28,29^ These models were estimated with the package frailtypack in R statistical software version 4.1.2 (2021-11-01, more details in the Supplementary Methods). The association between AV access and all-cause mortality, regardless of whether death was preceded or not by any hospitalization, was estimated with weighted Cox proportional hazard (PH) models. All analyses were based on the 36 imputed datasets and summarized according to Rubin and Schencker’s rules.^30^

#### Sensitivity analysis

Our analyses rely on the strong hypothesis that the risk of studied events in patients with first-line AV graft would have been the same as that in patients with first-line AV fistula, had the former group received a fistula instead, conditionally on propensity scores weighting (conditional exchangeability).^31^ To check whether there was evidence of violation of this hypothesis, we assessed the balance in the weighted frequency of preoperative imaging and number of hospitalizations in the two years preceding AV access creation (which were considered a proxy of pre-existing comorbidities included in the main analysis propensity score model) across graft and fistula groups. Because we found significant differences across studied groups with regards to these two covariates, we re-estimated propensity scores taking preoperative imaging (binary variable) and number of hospitalizations before AV access creation (fitted as a cubic B-spline with 2 interior knots) into account. Joint frailty models were then re-estimated with the new inverse probability weighting. Sensitivity analysis were stratified by diabetes status.

## Results

The study population included 18,625 patients who started hemodialysis with a catheter and without previous AV access creation (Figure 1). The mean age (standard deviation, SD) was 68 (±15) years in the 17,671 patients with first-line AV fistula. Two-thirds (65%) of them were male, 45% had diabetes, 26% heart failure, and 19% peripheral artery disease. In comparison with patients with first-line AV fistula, the 954 patients with first-line AV graft were older (mean age 68 versus 72 years, respectively), required mobility assistance more often (17 *versus* 21%), and underwent first AV access creation later (mean time from hemodialysis initiation to first access of 4.2 months, versus 3.4 months, Table 1 and Figure S2). Characteristics of patients included in or excluded from the analysis based on propensity score estimation is given in Table S1. Inverse probability weighting (IPW) resulted in covariate balance (absolute standardized difference [ASD] <10%) in the overall study population (Table 1) and across subgroups of age, sex, diabetes, heart failure, and peripheral artery disease (Table S2). Unbalance persisted for time to AV access creation in the subgroup aged <70.

**Figure 1.**
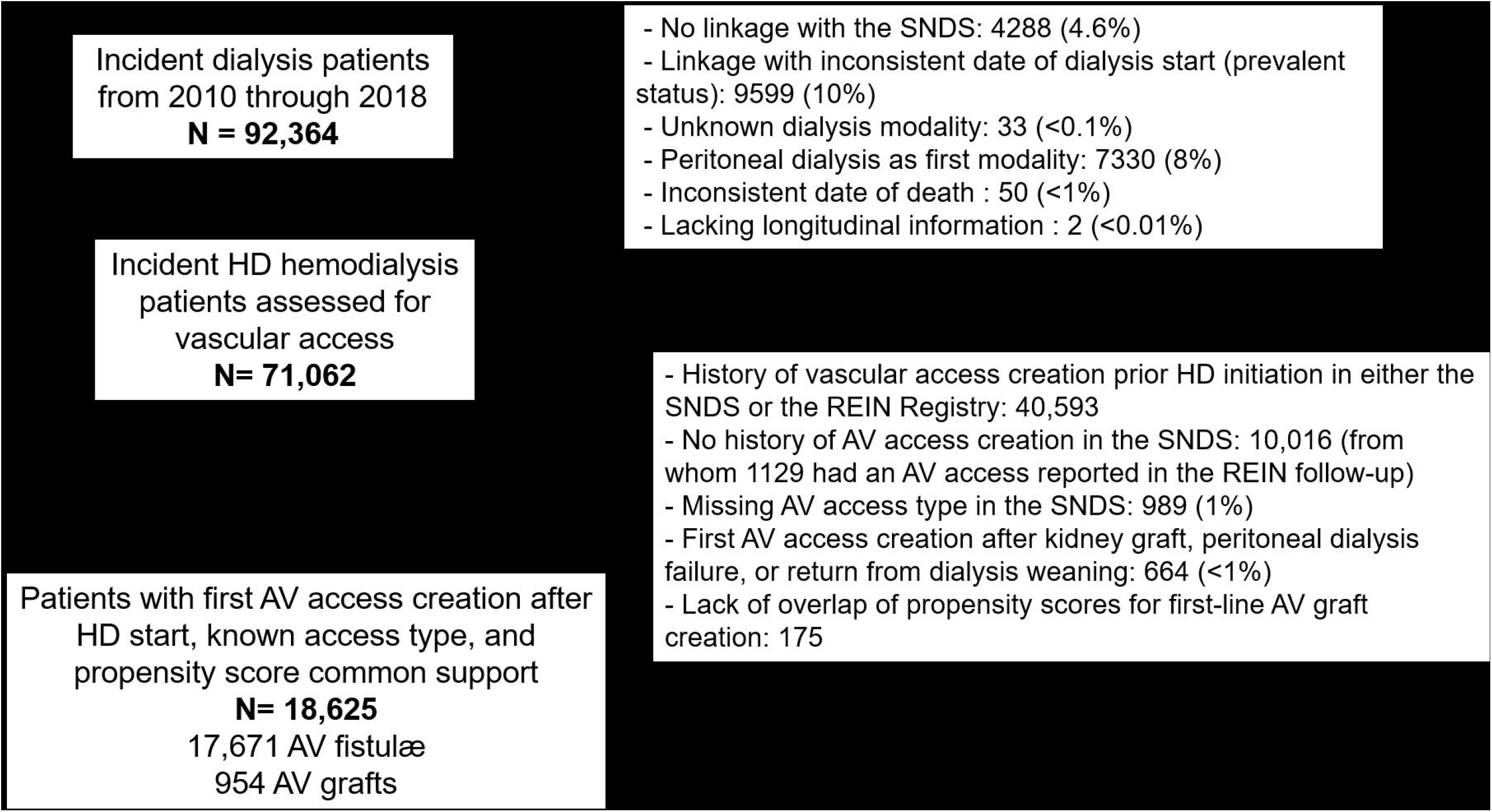
Selection of the study population. Abbreviations: AV, arteriovenous; HD, hemodialysis; SNDS, *Système National de Données de Santé*.

Over a median follow-up of 48 months (interquartile range [IQR] 27 - 48), 16,569 patients had at least one overnight hospital stay, for a total of 84,032 hospitalizations (median of 3 hospitalizations per patient, IQR 1-6), and 214,908 nights at hospital (median of 6 nights per patient, IQR 2-15); 6470 patients died. All-cause hospitalization rate for the overall study population was 1.99 per person-year (PY, Figure 2A). It was higher with older age (1.85 to 2.18 PY), and in the presence of diabetes (1.72 *versus* 2.31 PY), heart failure (1.86 *versus* 2.41 PY), or peripheral artery disease (1.84 *versus* 2.66 PY, Figure 2A). Overall, vascular access-cardiovascular-, and infection-related hospitalization rates were 0.41, 0.60, and 0.24 PY, respectively; subgroup patterns were mostly similar to that of all-cause hospitalization (Supplementary Figures 3A-C).

**Figure 2.**
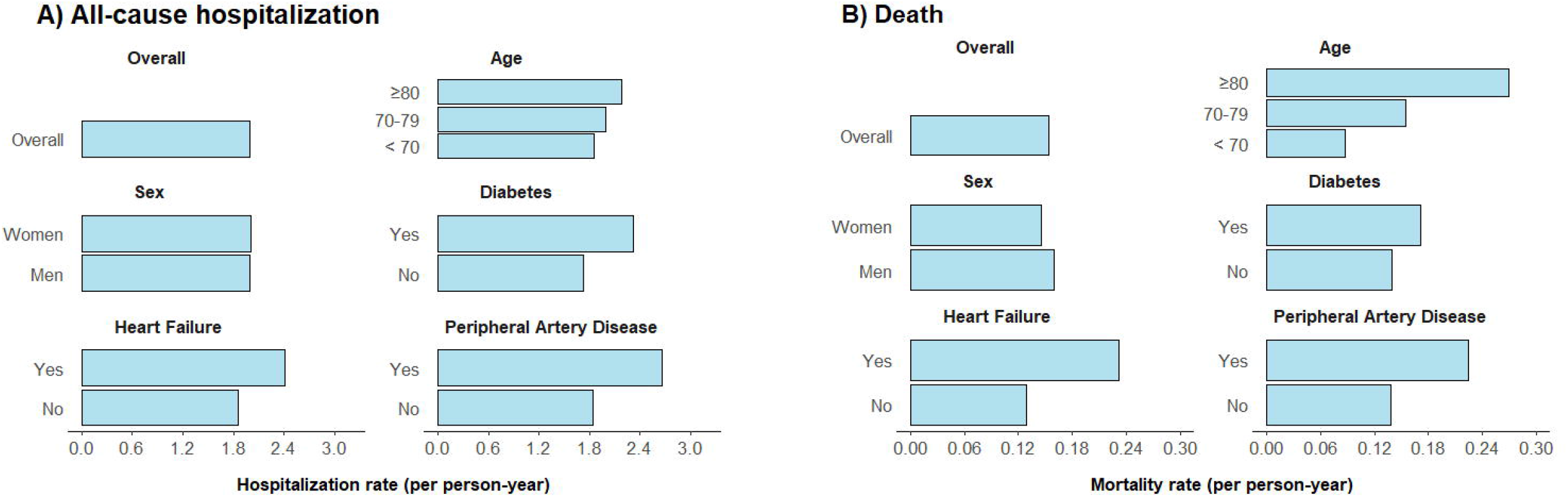
Crude all-cause hospitalization (A) and death (B) rates in the overall study population and across subgroups of age, sex, diabetes, heart failure, and peripheral artery disease.

Unweighted rates of all-cause and cause-specific hospitalizations (i.e., before IPW) were the highest among patients with first-line AV graft creation (Table 2). Median time to either first hospitalization or death was 8 (IQR 4-24) months for patients with first-line AV fistula and 6 (IQR 3-18) months for those with first-line AV graft (Figure 3). Patients with AV graft had a crude 14%-higher hazard of all-cause hospitalization (HR 1.14, 95% CI 1.08 to 1.20, Table 3), which was only slightly attenuated in IPW analysis (weighted wHR 1.11, 1.04 to 1.18). Associations of first-line AV graft with cardiovascular- and infection-related hospitalizations were of same magnitude (wHR 1.16, 1.01 to 1.33), but seemed stronger for vascular access-related hospitalizations (wHR 1.34, 1.21-1.49). First-line AV access type was associated with a higher, but not statistically significant, mortality hazard in Cox PH (wHR 1.09, 0.97-1.23), and the joint shared frailty model (in which correlation with all-cause hospitalization was accounted for, wHR 1.11, 0.77-1.61).

**Table 2.** Crude incidence (per person-year) of hospitalizations and all-cause death, by arteriovenous access type.

| Event | AV fistula |  | AV graft |  |
| --- | --- | --- | --- | --- |
|  | N events | Crude incidence (PY) | N events | Crude incidence (PY) |
| All-cause hospitalization | 78,929 | 1.97 | 4789 | 2.35 |
| Vascular access-related hospitalization | 16,245 | 0.41 | 1165 | 0.57 |
| Cardiovascular-related hospitalization | 23,869 | 0.60 | 1513 | 0.74 |
| Infection-related hospitalization | 9278 | 0.23 | 613 | 0.30 |
| All-cause death | 6054 | 0.15 | 412 | 0.20 |
| Before hospitalization | 787 | 0.07 | 60 | 0.13 |
| During or after a hospitalization | 2567 | 0.18 | 352 | 0.22 |

Hospitalization causes are not mutually exclusive. More than one cause of hospitalization may have been declared when the patient got transferred to another hospital. Hospitalization codes used are given in Supplementary box.
Abbreviations: AV, arteriovenous; PY, person-year.

**Table 3.** Hazard ratios (95% confidence intervals) of hospitalizations and death associated with first-line arteriovenous graft creation (versus fistula) in the overall population.

| Event type | Hazard ratio (95% CI) |  |
| --- | --- | --- |
|  | Unweighted | Weighted |
| **<u>Hospitalizations</u>** |  |  |
| All-cause | 1.14 (1.08-1.20) | 1.11 (1.04-1.18) |
| Vascular access-related | 1.42 (1.29-1.56) | 1.34 (1.21-1.49) |
| Cardiovascular-related | 1.31 (1.01-1.70) | 1.16 (1.05-1.29) |
| Infection-related | 1.24 (1.10-1.40) | 1.16 (1.01-1.33) |
| **<u>All-cause death</u>** |  |  |
| Cox PH model | 1.33 (1.20-1.47) | 1.09 (0.97-1.23) |
| Joint frailty model (accounting for the correlation between death and all-cause hospitalization) | 1.03 (0.89-1.19) | 1.11 (0.77-1.61) |

Heterogeneity across individuals (variance of the frailties) was significant in all models, as was the positive correlation between hospitalization and death.
Abbreviation: CI, confidence interval; PH, proportional hazard.

**Figure 3.**
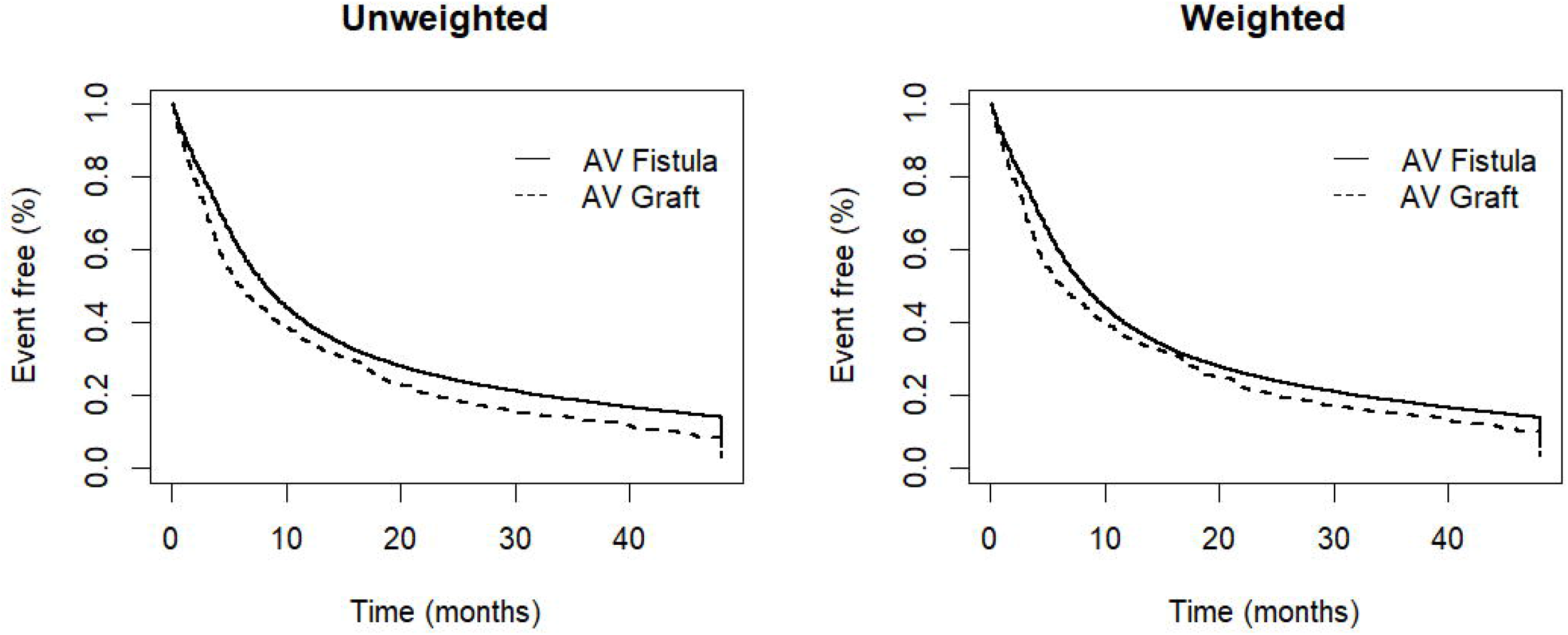
Unweighted and weighted event-free survival (all-cause hospitalization or death) by first-line arteriovenous access type. Weights correspond to the inverse probability of having one’s first-line arteriovenous access creation (inverse probability weighting). These probabilities were estimated with logistic regression, including as covariates: age, sex, year of hemodialysis start, primary kidney disease, diabetes, heart failure, peripheral artery disease, coronary heart disease, dysrhythmias, active malignancy, respiratory disease, liver disease, behavior disorders, mobility status, body mass index, urgent and intensive care unit dialysis start, kidney transplant waitlist status, and timing of AV access. Abbreviations: AV, arteriovenous.

Results were consistent between age subgroups (Figure 4 and Tables S3 to S6). The largest discrepancies were seen between patients without and with diabetes. The hazard of all-cause hospitalizations was 19% higher, and that of infection-related hospitalization, 37% higher with a first-line AV graft compared to fistula in patients without diabetes, while it did not differ between AV access groups in patients with diabetes. The weighted HR for hospitalizations due to AV access associated with first-line AV graft was also more prominent in patients without diabetes than with diabetes (wHR 1.53, 1.32-1.77, versus 1.17, 1.01-1.36, respectively), and in patients without heart failure than with heart failure (wHR 1.43, 1.26-1.62, and 1.15, 0.96-1.37). First-line AV graft, compared to fistula, was associated with higher hazard of infection-related hospitalization in women (wHR 1.45, 1.17-1.78), but not in men (wHR 1.02, 0.86-1.22). Weighted HR of death associated with first-line AV access, estimated with Cox PH models, were consistent in most subgroups, but differed across patients without (wHR 1.16, 1.00-1.34) and with peripheral artery disease (wHR 0.88, 0.70-1.09, Table S7). Joint frailty models showed significant positive correlation across hospitalizations within patients (nonzero frailty term) and between most hospitalization causes and death (alpha term >0, Tables S3 to S6).

**Figure 4.**
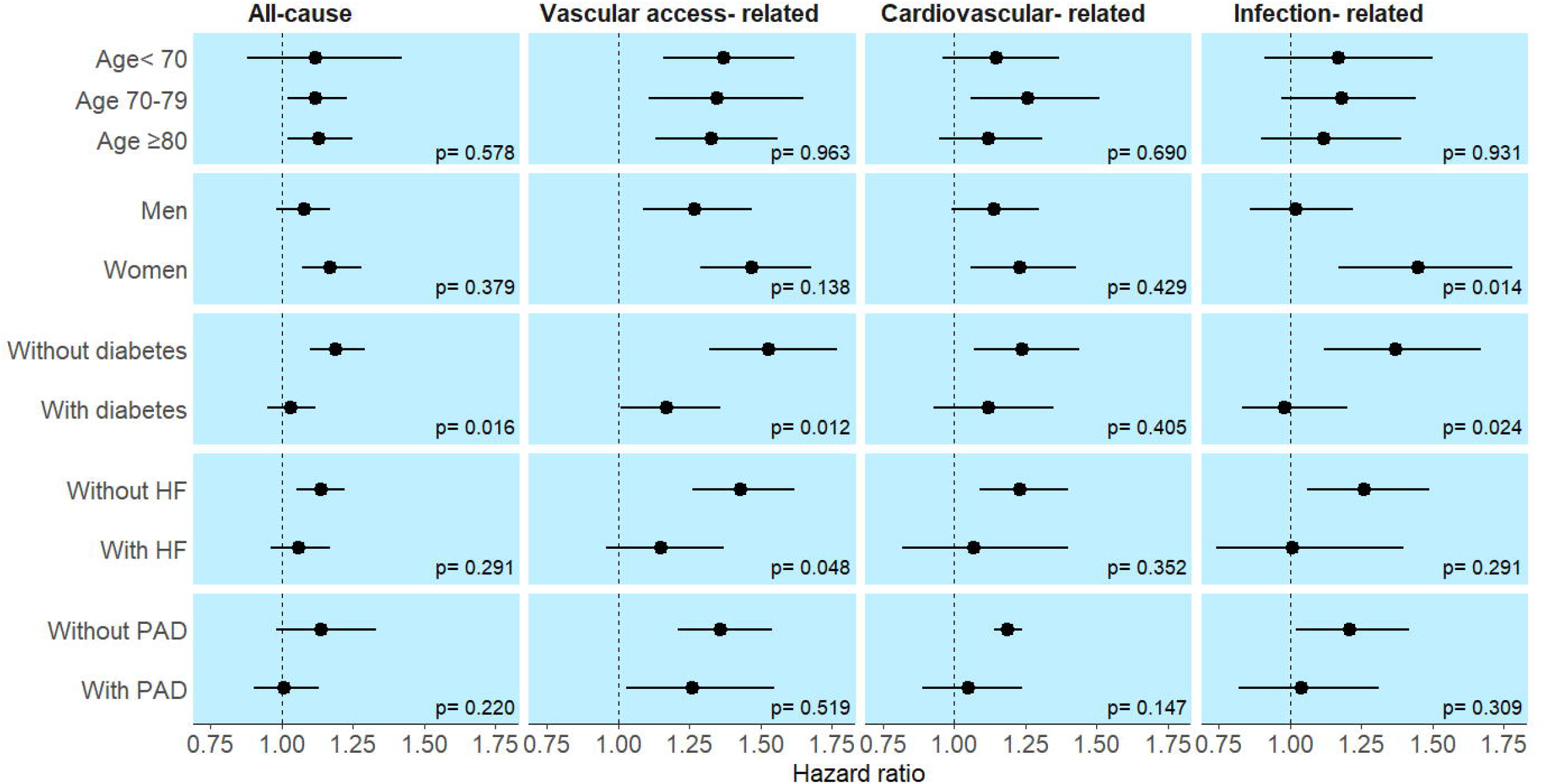
Weighted hazard ratios (95% confidence intervals) of hospitalizations associated with first-line arteriovenous graft creation (versus fistula), by subgroup. Weights correspond to the inverse probability of having one’s first-line arteriovenous access creation (inverse probability weighting). These probabilities were estimated with logistic regression, including as covariates: age, sex, year of hemodialysis start, primary kidney disease, diabetes, heart failure, peripheral artery disease, coronary heart disease, dysrhythmias, active malignancy, respiratory disease, liver disease, behavior disorders, mobility status, body mass index, urgent and intensive care unit dialysis start, kidney transplant waitlist status, and timing of AV access. Abbreviations: HF, heart failure; PAD, peripheral artery disease. Hazard ratios of hospitalizations were estimated with joint weighted semiparametric frailty models. Each joint model included a submodel for the hazard of recurrent hospitalizations (all cause or cause-specific hospitalizations) and a submodel for the hazard of all cause death, which was considered as informative censoring.

The weighted frequency of preoperative imaging and the mean number of hospitalizations in the two-year period preceding AV creation were higher among patients with first-line AV graft than in those with first-line AV fistula, regardless of diabetes status (Table S8). Sensitivity analysis in which propensity scores were re-estimated including this supplemental information showed slightly lower hazard ratios of hospitalizations associated with first-line AV graft than in the main analysis, but the direction and the difference in effect sizes between patients with and without diabetes were virtually unchanged (Table 4).

**Table 4.** Sensitivity analysis: Hazard ratios (95% confidence intervals) of hospitalizations associated with first-line arteriovenous graft creation (versus fistula) with re-estimated propensity score weighting, by diabetes status.

|  | Hazard ratio (95% confidence interval) |  |  |
| --- | --- | --- | --- |
|  | Unweighted | Weighted<br>(main analysis PS) | Weighted<br>(sensitivity analysis PS) |
| **<u>Without diabetes</u>** |  |  |  |
| All-cause hospitalization | 1.20 (1.11-1.29) | 1.19 (1.10-1.29) | 1.17 (1.06-1.28) |
| Vascular access-related hospitalization | 1.56 (1.36-1.78) | 1.53 (1.32-1.77) | 1.46 (1.25-1.70) |
| Cardiovascular-related | 1.31 (1.14-1.49) | 1.24 (1.07-1.44) | 1.20 (1.03-1.40) |
| Infection-related | 1.39 (1.16-1.67) | 1.45 (1.17-1.78) | 1.35 (1.09-1.67) |
| **<u>With diabetes</u>** |  |  |  |
| All-cause | 1.07 (0.99-1.15) | 1.03 (0.95-1.12) | 1.01 (0.92-1.10) |
| Vascular access-related | 1.28 (1.13-1.46) | 1.17 (1.01-1.36) | 1.15 (0.99-1.33) |
| Cardiovascular-related | 1.12 (0.99-1.26) | 1.12 (0.93-1.35) | 1.13 (0.87-1.47) |
| Infection-related | 1.10 (0.93-1.29) | 0.98 (0.83-1.20) | 0.95 (0.79-1.14) |

Abbreviation: PS, propensity score.

## Discussion

In this nationwide study including more than 18,000 patients who started hemodialysis with a catheter and underwent their first AV access creation after hemodialysis initiation, first-line AV graft was associated with increased risk of all-cause and cause-specific hospitalization, compared with AV fistula. Associations between first-line AV graft and hospitalization risk were consistent across age strata, but weaker and mostly not significant among patients with diabetes, compared with patients without this comorbidity. Association between first-line AV graft and all-cause death was marginal and not significant in most subgroups. While there was evidence of residual confounding in the main analysis, with unbalance in the frequency of preoperative imaging and in the mean number of hospitalizations prior AV access creation across study groups, sensitivity analysis taking this supplemental information into account led to the same conclusions. In light of the increasing amount of evidence calling into question the preference for AV fistula, our findings give additional insights about the influence of patients’ characteristics in vascular access-related morbidity and mortality.

Providing long-term care for kidney failure requires anticipating choices and complications leading to transitions across KRT modalities and across dialysis accesses. This process will depend on patient characteristics, such as age and comorbidity burden, but also patient expectations with regards to treatment. With this in view, the latest KDOQI (Kidney Disease Outcome and Quality Initiative) guidelines on vascular access has shifted from a focus on increasing AV fistula use (*Fistula First*) to a more patient-centered approach, the “*Patient Life-Plan first*”. KDOQI guidelines have also downgraded the level of evidence once assigned to the recommendation of AV fistula as preferred hemodialysis access, from moderately strong to low quality evidence. Guideline authors argue that previous observational studies which showed better outcomes with AV fistula versus AV graft or catheter were mostly based on the vascular access actually used (as opposed to that intended), thus ignoring complications that may arise during the maturation period and being more prompt to selection bias.

In this study, we assessed outcomes associated with first-line AV access, i.e. the first AV access created. We focused on patients who initiated hemodialysis with a catheter, and may have increased risk of fistula maturation failure. Overall, patients who received a first-line AV graft had higher rates of hospitalization requiring at least one overnight stay than those who received a fistula. However, the strength of this association seemed to vary across hospitalization types and to depend on comorbidity status. Overall, patients with first-line AV graft had 11% higher hazard of all-cause hospitalizations than their counterparts with first-line AV fistula. As expected, AV access type seemed more closely associated with vascular access-related hospitalizations (34% increased hazard) than with other cause-specific hospitalizations (16%). Of special note, first-line AV graft in patients with diabetes was not associated with increased hazard of either all-cause, or infection-related hospitalizations, and was associated with a less pronounced increased hazard of vascular access-related hospitalizations than in patients without diabetes (17% versus 53% respectively, *P* for interaction 0.012). The same pattern was observed across patients with or without heart failure and with or without peripheral artery disease, but the interaction term was not statistically significant in most of these analyses. In contrast with our hypothesis that first-line AV graft would more often prevent catheter use in women, first-line AV graft was significantly associated with increased hazard of infection related-hospitalizations in women, but not in men. The reasons for this finding are not clear and deserve further investigation. All-cause death was marginally and not significantly associated with AV access type in all groups but in patients without peripheral artery disease.

Our findings from the French REIN Registry partially contrast with the results from previous studies based on the United States Renal Data System (USRD). In incident hemodialysis patients from 2005 to 2008, De Silva et al^20^ first showed an absence of association between first-line AV access type and mortality. Stratified analyses in that study suggested effect modification by age, with equivalent survival rates across AV access groups being limited to patients older than 80. In a more recent USRDS cohort (2013 to 2014), Lyu et al^11^ reported similar survival and hospitalization rates in patients having received either a fistula or a graft as first-line AV access. In that study, however, first-line AV fistula remained associated with 24 to 27% lower hazard of vascular access-related hospitalization after taking account of potential confounders with, respectively, instrumental variable analysis and propensity score weighting.

Several reasons may explain the differences between findings from those studies and ours. First, USRDS-based studies,^20,21^ while also focused on patients starting hemodialysis with catheter without previous AV access placement, were restricted to patients aged ≥67 years. Mean age in patients with fistula and graft was thus much higher in the study from Lyu et al^21^ (76 and 78 years, respectively) then in ours (68 and 72 years), as was their population frequency of comorbidities. Interestingly, despite these differences, rates of all-cause and most cause-specific hospitalization reported by that American study were similar to ours (requiring at least one hospital overnight stay). All-cause hospitalization rate – which do not depend on hospitalization codes retained for analysis – was 1.73 and 2.11 per patient-year in patients with first-line AV fistula and graft in the American study, respectively, *versus* 1.97 and 2.35 in our study. Even more striking was the difference in the rate of vascular access-related hospitalizations: 0.09 and 0.17 per patient-year in patients with first-line AV fistula and graft in the American study, respectively, *versus* 0.41 and 0.57 in ours. Thus, besides differences in population characteristics, these data suggest that variation in clinical practices may play an important role in findings regarding performance of vascular access in different settings.

The DOPPS (Dialysis Outcomes and Practice Patterns Study) has, since the early 2000’s, highlighted international variations in practices and outcomes related with hemodialysis vascular access, ^32^ although international data on patterns of AV graft creation and use are still scarce. With regards to AV fistula, DOPPS reported substantial variation in the location of this access in patients recruited from national samples of randomly select hemodialysis facilities, with 68% of AV fistula being created in the upper arm in the United States, *versus* 34-47% in Europe/ Australia-New Zealand, and 5-15% in Japan.^33,34^ Upper arm AV accesses, in addition to affect future vascular access options, have been associated with a higher frequency of steal syndrome,^35^ and of complications from high-flow AV access.^36^ Surgical training may also be a relevant aspect in AV access outcomes. A lower volume of previous AV fistula creation at the surgeon- or the provider-level has been associated with a lower likelihood of AV fistula maturating requiring prolonged catheter use.^39,40^ In addition to AV access type, these and other aspects related to AV access creation, use, and management of complications are certainly critical for AV access outcomes but are beyond the scope of this study.

Our study strengths include its large, registry-based population; the granularity of the data regarding AV access procedures and hospitalizations provided by the linkage with the French national health-administrative database; the use of information on AV access creation, rather than that on AV access use, to define study groups; consideration of pre-specified subgroup analysis in the construction of the propensity score; and the use of joint frailty models, which appropriately account for event recurrence and informative censoring by death. Our study has also limitations. Linkage of the REIN Registry with the SNDS was based on an indirect, deterministic algorithm, which is subject to mismatch. To deal with this limitation, we restricted our analysis to patients whose linkage was based on information with the highest degree of reliability (date of dialysis start, and of kidney transplant or death when available), and who had consistent AV access information according to the two databases. Because the SNDS is designed for reimbursement purposes, there may exist some misclassification of AV access types and hospitalizations, but which are likely to be non-differential (independent from study groups and outcomes). Despite the use of propensity score weighting, which included a number of covariates related with AV access choice, we cannot rule out our results being at least partly explained by confounding bias. AV access groups remained unbalanced after propensity score weighting with regards to the frequency of preoperative imaging and rates of hospitalizations in the two years preceding AV access creation (which were not accounted for in propensity score weighting of the main analysis). Nevertheless, when we included this information in the propensity score, results were virtually unchanged. It is worth noting that differences in the number of hospitalizations prior AV access creation across study groups were of the same relative magnitude in patients with or without diabetes, and thus are not likely to explain differences in the relation between first-line AV graft and study outcomes according to diabetes status.

In conclusion, in patients starting hemodialysis with a catheter without previous AV access creation in France, the first-line AV graft was associated with a higher hazard of hospitalization. This may, however, not be the case for patients with a poor vascular condition, who had a similar rates of hospitalization and mortality with either first-line AV graft or fistula. In the absence of increased hospital morbidity and mortality with AV graft in this subpopulation, outcomes related with more timely AV graft creation, use and patient preferences deserve to be further explored by future research.

## Data Availability

All data used in the present study are available upon reasonable request to the REIN scientific committee at the French Agence of Biomedecine.

## Article information

### Authors’ contribution

Research idea and study design: NAP, KL; data acquisition: CC, FG, MPR; data analysis/interpretation: NAP, MPR, JH, CC, FG, CC, VR, KL; statistical analysis: NAP; supervision or mentorship: VR, KL. Each author contributed important intellectual content during manuscript drafting or revision and agrees to be personally accountable for the individual’s own contributions and to ensure that questions pertaining to the accuracy or integrity of any portion of the work, even one in which the author was not directly involved, are appropriately investigated and resolved, including with documentation in the literature if appropriate.

### Support

This work was supported by a research grant from the French Biomedicine Agency, Recherche REIN 2019, number 19REIN001.

## Acknowledgments

The authors thank all the participants at the REIN Registry and the Agence de la Biomédecine, especially the nephrologists and professionals responsible for data collection and quality control. Dialysis centers participating in the registry are listed in the REIN annual report (https://www.agence-biomedecine.fr/IMG/pdf/rapport_rein_2019_2021-10-14.pdf).

## Supplementary online material

### Supplemental methods

Supplementary Box. Main hospitalization diagnosis (DGN_PAL), according to the International Classification of Diseases ICD-10, and the French diagnosis related groups (GRG_GHM)* used to identify cause-specific hospitalization.

Table S1. Characteristics of patients included in or excluded from the analysis based on propensity score common support (corresponding to the overlap of propensity scores).

Table S2. Assessment of covariate balance# across AV access groups through the 36 imputed datasets, overall and by subgroups.

Table S3. Hazard ratios (95% confidence intervals) of all-cause hospitalization associated with first-line AV graft creation (versus AV fistula), overall and by subgroup.

Table S4. Hazard ratios (95% confidence intervals) of vascular access-related hospitalization associated with first-line AV graft creation (versus AV fistula), overall and by subgroup.

Table S5. Hazard ratios (95% confidence intervals) of cardiovascular-related hospitalization associated with first-line AV graft creation (versus AV fistula), overall and by subgroup.

Table S6. Hazard ratios (95% confidence intervals) of infection-related associated with first-line AV graft creation (versus AV fistula), overall and by subgroup.

Table S7. Hazard ratios (95% confidence intervals) of all-cause death associated with first-line AV graft creation (versus AV fistula), overall and by subgroup.

Table S8. Sensitivity analysis: Assessment of the balance in the frequency of preoperative imaging and the number of hospitalizations in the two years preceding arteriovenous access creation, by diabetes status.

Figure S1. Direct acyclic graph for the relation between first-line arteriovenous access and outcomes.

Figure S2. Unweighted and weighted distributions of time from dialysis start to arteriovenous access creation.

Figure S3. Vascular access- (A), cardiovascular- (B), and infection-related (C) hospitalization rates in the overall study population and across subgroups of age, sex, diabetes, heart failure, and peripheral artery disease.

