## Supplementary online material for "First-line arteriovenous access and risks of hospitalization and death in patients starting hemodialysis – a nationwide cohort study"

### Supplemental methods

### Propensity score model

In the propensity score model, age, BMI, and time of VA access creation were modeled with natural cubic splines with 4, 3, and 2 knots placed at the percentiles, respectively. The number of knots was chosen based on the AIC criterion. An interaction term between sex and BMI was included in the model to improve covariate balance between the two AV access groups (fistula or graft). To reduce the potential violation of the positivity assumption inherent to causal inference with propensity scores (which requires that patients in each group has a non-null probability to receive the other AV access),^26^ only patients in the common support (corresponding to the overlap of propensity scores) were included in the analysis.^28^ Characteristics of patients included in or excluded from the analysis were compared (Table S1). Finally, propensity scores were re-estimated among included patients only as recommended by Sturmer et al.^28^ Covariate balance was assessed in the overall sample and in each subgroup.

### Joint weighted semiparametric frailty models

Shared random effects accounted for the dependence between recurrent hospitalizations and death. Penalized weighted likelihood was used to avoid parametric assumption on baseline hazards^30^ . We used the calendar timescale for the recurrent times of hospital discharges, i.e. the time elapsed since the beginning of the study (AV access creation) until the time of hospitalization. However, a patient is considered at risk of a jth hospitalization only after the (j-1)th hospitalization. We checked the proportional hazard assumption by comparing the approximated likelihood cross-validation criterion (the LCV, equivalent of the Akaïke criterion in the semiparametric case) of our models to those of alternative models including a time varying effect of AV graft (a linear combination of cubic B-splines with 2, 3 and 4 interior knots).

### Supplementary Box. Main hospitalization diagnosis (DGN_PAL), according to the International Classification of Diseases ICD-10, and the French diagnosis related groups (GRG_GHM)* used to identify cause-specific hospitalization.

| Type of hospitalization | Codes |
| --- | --- |
| Vascular access- related | GRG_GHM like "11C09%" or GRG_GHM like "05C21%" or GRG_GHM like "05K26%" or GRG_GHM like "11K07Z" or GRG_GHM like "05K14Z" or GRG_GHM like "24C39Z" or GRG_GHM like "24K04Z" or    DGN_PAL like "T823" or DGN_PAL like "I770" or DGN_PAL like "T824" or DGN_PAL like "T825" or  DGN_PAL like "T827" or DGN_PAL like "T828" or DGN_PAL like "T856" or DGN_PAL like "T857" or DGN_PAL like "Z452" |
| Cardiovascular- related | DGN_PAL like "I48%" or DGN_PAL like "I10%" or DGN_PAL like "I11%" or DGN_PAL like "I12%" or DGN_PAL like "I13%" or DGN_PAL like "I14%" or DGN_PAL like "I15%" or DGN_PAL like "I16%" or DGN_PAL like "I21%" or DGN_PAL like "I22%" or DGN_PAL like "I252" or DGN_PAL like "I099" or DGN_PAL like "I110" or DGN_PAL like "I130" or DGN_PAL like "I132" or DGN_PAL like "I255" or DGN_PAL like "I420" or DGN_PAL like "I425" or DGN_PAL like "I426" or DGN_PAL like "I427" or DGN_PAL like "I428" or DGN_PAL like "I429" or DGN_PAL like "I43%" or DGN_PAL like "I50%" or DGN_PAL like "I70%" or DGN_PAL like "I71%" or DGN_PAL like "I731" or DGN_PAL like "I738" or DGN_PAL like "I739" or DGN_PAL like "I771" or DGN_PAL like "I790" or DGN_PAL like "I792" or DGN_PAL like "Z958" or DGN_PAL like "Z959" or DGN_PAL like "E105" or DGN_PAL like "E115" or DGN_PAL like "E135" or DGN_PAL like "G45%" or DGN_PAL like "G46%" or DGN_PAL like "H340" or DGN_PAL like "I6%" or DGN_PAL like "I05%" or DGN_PAL like "I06%" or DGN_PAL like "I07%" or DGN_PAL like "I08%" or DGN_PAL like "I090" or  DGN_PAL like "I091" or DGN_PAL like "I098" or DGN_PAL like "I1%" or DGN_PAL like "I2%" or DGN_PAL like "I31%" or DGN_PAL like "I32%" or DGN_PAL like "I339" or DGN_PAL like "I34%" or DGN_PAL like "I35%" or DGN_PAL like "I36%" or DGN_PAL like "I37%" or DGN_PAL like "I38%" or DGN_PAL like "I401" or DGN_PAL like "I409" or DGN_PAL like "I42%" or DGN_PAL like "I43%" or DGN_PAL like "I44%" or DGN_PAL like "I45%" or DGN_PAL like "I46%" or DGN_PAL like "I47%" or DGN_PAL like "I48%" or DGN_PAL like "I49%" or DGN_PAL like "I5%" or DGN_PAL like "I6%" or DGN_PAL like "I7%" or DGN_PAL like "I80%" or DGN_PAL like "I81%" or DGN_PAL like "I82%" or DGN_PAL like "I83%" or DGN_PAL like "I84%" or DGN_PAL like "I85%" or DGN_PAL like "I86%" or DGN_PAL like "I87%" or DGN_PAL like "I89%" or DGN_PAL like "I95%" or  DGN_PAL like "I96%" or DGN_PAL like "I970" or DGN_PAL like "I971" or DGN_PAL like "I972" or DGN_PAL like "I998" or DGN_PAL like "I999" or DGN_PAL like "K64%" or DGN_PAL like "M30%" or DGN_PAL like "M31%" or DGN_PAL like "M321" or DGN_PAL like "N262" or DGN_PAL like "R000" or DGN_PAL like "R58%" or DGN_PAL like "T800" or DGN_PAL like "T817" or DGN_PAL like "T828" |
| Infection- related | GRG_GHM like "18M07%" or  DGN_PAL like "A40%" or DGN_PAL like "A41%" or DGN_PAL like "A0%" or DGN_PAL like "A1%" or DGN_PAL like "A2%" or DGN_PAL like "A31%" or DGN_PAL like "A32%" or DGN_PAL like "A35%" or DGN_PAL like "A36%" or DGN_PAL like "A37%" or DGN_PAL like "A38%" or DGN_PAL like "A39%" or DGN_PAL like "A4%" or DGN_PAL like "A5%" or DGN_PAL like "A6%" or DGN_PAL like "A7%" or DGN_PAL like "A8%" or DGN_PAL like "A9%" or DGN_PAL like "B%" or DGN_PAL like "D86%" or DGN_PAL like "E321" or DGN_PAL like "E832" or DGN_PAL like "G00%" or DGN_PAL like "G01%" or DGN_PAL like "G02%" or DGN_PAL like "G03%" or DGN_PAL like "G040" or DGN_PAL like "G042" or DGN_PAL like "G043" or DGN_PAL like "G048" or DGN_PAL like "G049" or DGN_PAL like "G05%" or DGN_PAL like "G06%" or DGN_PAL like "G07%" or DGN_PAL like "G08%" or DGN_PAL like "G09%" or DGN_PAL like "G14%" or DGN_PAL like "G374" or DGN_PAL like "G92%" or DGN_PAL like "G937" or DGN_PAL like "H0%" or DGN_PAL like "H10%" or DGN_PAL like "H162" or DGN_PAL like "H32%" or DGN_PAL like "H660" or DGN_PAL like "H661" or DGN_PAL like "H662" or DGN_PAL like "H663" or DGN_PAL like "H66%" or DGN_PAL like "H67%" or DGN_PAL like "H70%" or DGN_PAL like "H75%" or DGN_PAL like "H830" or DGN_PAL like "H921" or DGN_PAL like "H950" or DGN_PAL like "H951" or DGN_PAL like "I00%" or DGN_PAL like "I01%" or DGN_PAL like "I02%" or DGN_PAL like "I092" or DGN_PAL like "I32%" or DGN_PAL like "I330" or DGN_PAL like "H830" or DGN_PAL like "I39%" or DGN_PAL like "I40%" or DGN_PAL like "I41%" or DGN_PAL like "I673" or DGN_PAL like "J0%" or DGN_PAL like "J10%" or DGN_PAL like "J11%" or DGN_PAL like "J12%" or DGN_PAL like "J13%" or DGN_PAL like "J14%" or DGN_PAL like "J15%" or DGN_PAL like "J16%" or DGN_PAL like "J17%" or DGN_PAL like "J181" or DGN_PAL like "J188" or DGN_PAL like "J189" or DGN_PAL like "J19%" or DGN_PAL like "J20%" or DGN_PAL like "J21%" or DGN_PAL like "J31%" or DGN_PAL like "J32%" or DGN_PAL like "J350" or DGN_PAL like "J36%" or DGN_PAL like "J370" or DGN_PAL like "J371" or DGN_PAL like "J390" or DGN_PAL like "J391" or DGN_PAL like "J392" or DGN_PAL like "J40%" or DGN_PAL like "J411" or DGN_PAL like "J47%" or DGN_PAL like "J371" or DGN_PAL like "J850" or DGN_PAL like "J851" or DGN_PAL like "J852" or DGN_PAL like "J86%" or DGN_PAL like "J87%" or DGN_PAL like "J88%" or DGN_PAL like "J89%" or DGN_PAL like "J90%" or DGN_PAL like "J91%" or DGN_PAL like "J92%" or DGN_PAL like "J94%" or DGN_PAL like "K046" or DGN_PAL like "K047" or DGN_PAL like "K113%" or DGN_PAL like "K122" or DGN_PAL like "K352" or DGN_PAL like "K353" or DGN_PAL like "K358" or DGN_PAL like "K36%" or DGN_PAL like "K37%" or DGN_PAL like "K51%" or DGN_PAL like "K570" or DGN_PAL like "K572" or DGN_PAL like "K574" or DGN_PAL like "K578" or DGN_PAL like "K61%" or DGN_PAL like "K630" or DGN_PAL like "K65%" or DGN_PAL like "K67%" or DGN_PAL like "K68%" or DGN_PAL like "K71%" or DGN_PAL like "K750" or DGN_PAL like "K751" or DGN_PAL like "K752" or DGN_PAL like "K753" or DGN_PAL like "K758" or DGN_PAL like "K759" or DGN_PAL like "K764" or DGN_PAL like "K77%" or DGN_PAL like "K81%" or DGN_PAL like "L01%" or DGN_PAL like "L02%" or DGN_PAL like "L03%" or DGN_PAL like "L04%" or DGN_PAL like "L05%" or DGN_PAL like "L06%" or DGN_PAL like "L07%" or DGN_PAL like "L08%" or DGN_PAL like "L444" or DGN_PAL like "L702" or DGN_PAL like "L88%" or DGN_PAL like "L928" or DGN_PAL like "L946" or DGN_PAL like "L980" or DGN_PAL like "L983" or DGN_PAL like "M00%" or DGN_PAL like "M01%" or DGN_PAL like "M021" or DGN_PAL like "M023" or DGN_PAL like "M028" or DGN_PAL like "M652" or DGN_PAL like "M462" or DGN_PAL like "M463" or DGN_PAL like "M86%" or DGN_PAL like "M908" or DGN_PAL like "N10%" or DGN_PAL like "N11%" or DGN_PAL like "N12%" or DGN_PAL like "N136" or DGN_PAL like "N151" or DGN_PAL like "N159" or DGN_PAL like "N16%" or DGN_PAL like "N300" or DGN_PAL like "N303" or DGN_PAL like "N308" or DGN_PAL like "N340" or DGN_PAL like "N341" or DGN_PAL like "N342" or DGN_PAL like "N343" or DGN_PAL like "N351" or DGN_PAL like "N37%" or DGN_PAL like "N390" or DGN_PAL like "N41%" or DGN_PAL like "N45%" or DGN_PAL like "N481" or DGN_PAL like "N482" or DGN_PAL like "N49%" or DGN_PAL like "N51%" or DGN_PAL like "N61%" or DGN_PAL like "N70%" or DGN_PAL like "N71%" or DGN_PAL like "N72%" or DGN_PAL like "N73%" or DGN_PAL like "N74%" or DGN_PAL like "N751" or DGN_PAL like "N751" or DGN_PAL like "N760" or DGN_PAL like "N761" or DGN_PAL like "N762" or DGN_PAL like "N763" or DGN_PAL like "N764" or DGN_PAL like "N771" or DGN_PAL like "N980" or DGN_PAL like "O85%" or DGN_PAL like "O86%" or DGN_PAL like "R091" or DGN_PAL like "R091" or DGN_PAL like "T802" or DGN_PAL like "T814" or DGN_PAL like "T826" or DGN_PAL like "T827" or DGN_PAL like "T835" or DGN_PAL like "T836" or DGN_PAL like "T845" or DGN_PAL like "T846" or DGN_PAL like "T847" or DGN_PAL like "T874" or DGN_PAL like "T880" |

* The French diagnosis related groups (GHM) classifies all hospital care that are related with each order from a medico-economic perspective. It is generated by an algorithm of the French social security based on information from the hospital discharge summary for each patient. For more information about GHM, see [solidarites-sante.gouv.fr](https://solidarites-sante.gouv.fr/professionnels/gerer-un-etablissement-de-sante-medico-social/financement/financement-des-etablissements-de-sante-10795/financement-des-etablissements-de-sante-glossaire/article/groupe-homogene-de-malades-ghm)

### Table S1. Characteristics of patients included in or excluded from the analysis based on propensity score common support (corresponding to the overlap of propensity scores).

|  | Included | Excluded | Unweighted absolute standardized difference |
| --- | --- | --- | --- |
|  | N= 18,625 (99%) | N= 175 (1%) |  |
| Age, mean (SD) | 68 (15) | 42 (13) | 1.84 |
| Women, % | 35 | 1 | 0.34 |
| AV graft, % | 5 | 0.05 | 0.05 |
| Year of HD start, % |  |  |  |
| 2010 | 10 | 22 | 0.12 |
| 2011 | 10 | 8 | 0.02 |
| 2012 | 11 | 40 | 0.30 |
| 2013 | 11 | 8 | 0.03 |
| 2014 | 11 | 2 | 0.10 |
| 2015 | 12 | 2 | 0.09 |
| 2016 | 12 | 1 | 0.11 |
| 2017 | 12 | 8 | 0.04 |
| 2018 | 11 | 8 | 0.03 |
| Primary kidney disease, % |  |  |  |
| Diabetes | 24 | 4 | 0.20 |
| Hypertension or vascular disease | 26 | 9 | 0.17 |
| Glomerulonephritis | 11 | 23 | 0.12 |
| Pyelonephritis | 5 | 5 | 0.003 |
| Polycystic kidney disease | 2 | 11 | 0.09 |
| Other | 17 | 31 | 0.14 |
| Unknown | 16 | 17 | 0.02 |
| Diabetes, % | 45 | 16 | 0.30 |
| Heart failure, % | 17 | 4 | 0.23 |
| Peripheral artery disease, % | 20 | 1 | 0.18 |
| Dysrhythmia, % | 23 | 5 | 0.18 |
| Cerebrovascular disease, % | 11 | 0.5 | 0.10 |
| Coronary heart disease, % | 26 | 3 | 0.22 |
| Respiratory disease, % | 16 | 4 | 0.12 |
| Liver disease, % | 5 | 0.3 | 0.05 |
| Active malignancy, % | 12 | 6 | 0.07 |
| Behavior disorders, % | 3 | 0.8 | 0.02 |
| Mobility status (partially or totally dependent), % | 17 | 1 | 0.16 |
| Body mass index, mean (SD) | 26 (6) | 31 (5) | 0.91 |
| Urgent dialysis start | 53 | 76 | 0.23 |
| ICU dialysis start | 17 | 10 | 0.06 |
| Waitlisted for KTx | 3 | 0.2 | 0.03 |
| Time (months) from dialysis start to AV access creation, mean (SD) | 3.4 (4.5) | 4.2 (6.2) | 0.18 |

Weights correspond to the inverse probability of having one’s first-line arteriovenous access creation (inverse probability weighting). These probabilities were estimated with logistic regression, including as covariates: age, sex, year of hemodialysis start, primary kidney disease, diabetes, heart failure, peripheral artery disease, coronary heart disease, dysrhythmias, active malignancy, respiratory disease, liver disease, behavior disorders, mobility status, body mass index, urgent and intensive care unit dialysis start, kidney transplant waitlist status, and timing of AV access. All statistics are based on the summary of the 36 imputed datasets, and the number of patients by AV access type is the median number in each group through these datasets.

Abbreviations: AV, arteriovenous; HD, hemodialysis; ICU, intensive care unit; KTx, kidney transplantation; SD, standard deviation

### Table S2. Assessment of covariate balance^#^ across AV access groups through the 36 imputed datasets, overall and by subgroups.

| Sample | Number of patients in the AV fistula group | Number of patients in the AV graft group | Number of balanced  datasets | Unbalanced covariates  (and number of datasets) | Min-max relative sample loss^¥^ in the AV graft group (%) |
| --- | --- | --- | --- | --- | --- |
| All patients | 17671 | 954 | 36 | - | 20-22 |
| Age < 70 | 8428 | 356 | 36 | Time of AV access placement (n=36) | 17-20 |
| Age 70-79 | 4593 | 273 | 0 | - | 16-18 |
| Age ≥ 80 | 4649 | 325 | 36 | - | 14-17 |
| Women | 6143 | 411 | 36 | - | 15-17 |
| Men | 11528 | 543 | 36 | - | 18-21 |
| Without diabetes | 9717 | 469 | 36 | - | 18-21 |
| With diabetes | 7954 | 485 | 36 | - | 19-21 |
| Without HF | 13030 | 638 | 36 | - | 18-21 |
| With HF | 4642 | 316 | 36 | - | 18-21 |
| Without PAD | 14260 | 709 | 36 | - | 18-21 |
| With PAD | 3410 | 245 | 35 | BMI (n=1) | 18-23 |

\# Assessment of covariate balance was performed with weighted absolute standardized difference of patient characteristics across AV access groups. Differences >0.1 were considered significant.

¥ Relative sample loss refers to the effective to real sample size ratio, where effective sample size is $\frac{{sum\left( weights \right)}^{2}}{sum\left( {weights}^{2} \right)}$. It is an indicator of variance increase due to weighting.

### Table S3. Hazard ratios (95% confidence intervals) of all-cause hospitalization associated with first-line AV graft creation (versus AV fistula), overall and by subgroup.

| Sample | All-cause hospitalization | | | | Variance of random effect* | Alpha* | *P* for interaction  (weighted HR) |
| --- | --- | --- | --- | --- | --- | --- | --- |
|  | Unweighted | | Weighted | |  |  |  |
| All patients | 1.14 (1.08-1.20) | | 1.11 (1.04-1.18) | | 0.83 (p<0.001) | 1.66 (p<0.001) | - |
| Age |  | |  | |  |  | 0.578 |
| < 70 | 1.14 (1.04-1.25) | | 1.12 (0.88-1.42) | | 0.79 (p<0.001) | 1.53 (p<0.001) |  |
| 70-79 | 1.11 (1.01-1.22) | | 1.12 (1.02-1.23) | | 0.78 (p<0.001) | 1.82 (p<0.001) |  |
| ≥ 80 | 1.17 (1.06-1.30) | | 1.13 (1.02-1.25) | | 0.99 (p<0.001) | 1.76 (p<0.001) |  |
| Sex |  | |  | |  |  | 0.379 |
| Women | 1.17 (1.08-1.27) | | 1.17 (1.07-1.28) | | 0.72 (p<0.001) | 1.60 (p<0.001) |  |
| Men | 1.11 (1.03-1.19) | | 1.08 (0.98-1.17) | | 0.90 (p<0.001) | 1.65 (p<0.001) |  |
| Diabetes |  | |  | |  |  | 0.016 |
| No | 1.20 (1.11-1.29) | | 1.19 (1.10-1.29) | | 0.92 (p<0.001) | 1.89 (p<0.001) |  |
| Yes | 1.07 (0.99-1.15) | | 1.03 (0.95-1.12) | | 0.72 (p<0.001) | 1.45 (p<0.001) |  |
| Heart failure |  | |  | |  |  | 0.291 |
| No | 1.16 (1.09-1.25) | | 1.14 (1.05-1.22) | | 0.83 (p<0.001) | 1.76 (p<0.001) |  |
| Yes | 1.07 (0.97-1.18) | | 1.06 (0.96-1.17) | | 0.76 (p<0.001) | 1.39 (p<0.001) |  |
| Peripheral artery disease | |  | |  |  |  | 0.220 |
| No | 1.17 (0.97-1.42) | | 1.14 (0.98-1.33) | | 0.85 (p<0.001) | 1.67 (p<0.001) |  |
| Yes | 1.02 (0.91-1.14) | | 1.01 (0.90-1.13) | | 0.66 (p<0.001) | 1.51 (p<0.001) |  |

Weights correspond to the inverse probability of having one’s first-line arteriovenous access creation (inverse probability weighting). These probabilities were estimated with logistic regression, including as covariates: age, sex, year of hemodialysis start, primary kidney disease, diabetes, heart failure, peripheral artery disease, coronary heart disease, dysrhythmias, active malignancy, respiratory disease, liver disease, behavior disorders, mobility status, body mass index, urgent and intensive care unit dialysis start, kidney transplant waitlist status, and timing of AV access.

Hazard ratios of hospitalizations were estimated with joint weighted semiparametric frailty models. Each joint model included a submodel for the hazard of recurrent hospitalizations (all cause or cause-specific hospitalizations) and a submodel for the hazard of all cause death, which was considered as informative censoring.

*****Median variance of random effect and alpha in the weighted model through 36 imputed datasets. Positives values of alpha indicate that all-cause hospitalization and death are positively correlated.

### Table S4. Hazard ratios (95% confidence intervals) of vascular access-related hospitalization associated with first-line AV graft creation (versus AV fistula), overall and by subgroup.

| Sample | Vascular access-related hospitalization | | | | Variance of random effect* | Alpha* | *P* for interaction  (weighted HR) |
| --- | --- | --- | --- | --- | --- | --- | --- |
|  | Unweighted | | Weighted | |  |  |  |
| All patients | 1.42 (1.29-1.56) | | 1.34 (1.21-1.49) | | 0.88 (p<0.001) | 0.08 (p<0.001) | - |
| Age |  | |  | |  |  | 0.963 |
| < 70 | 1.47 (1.26-1.71) | | 1.37 (1.16-1.62) | | 0.97 (p<0.001) | 0.17 (p<0.001) |  |
| 70-79 | 1.36 (1.13-1.63) | | 1.35 (1.11-1.65) | | 0.90 (p<0.001) | 0.00 (p=0.791) |  |
| ≥ 80 | 1.43 (1.23-1.67) | | 1.33 (1.13-1.56) | | 0.72 (p<0.001) | 0.11 (p<0.001) |  |
| Sex |  | |  | |  |  | 0.138 |
| Women | 1.45 (1.29-1.64) | | 1.47 (1.29-1.68) | | 0.64 (p<0.001) | 0.07 (p=0.034) |  |
| Men | 1.33 (1.13-1.56) | | 1.27 (1.09-1.47) | | 1.03 (p<0.001) | 0.13 (p<0.001) |  |
| Diabetes |  | |  | |  |  | 0.012 |
| No | 1.56 (1.36-1.78) | | 1.53 (1.32-1.77) | | 0.86 (p<0.001) | 0.15 (p<0.001) |  |
| Yes | 1.28 (1.13-1.46) | | 1.17 (1.01-1.36) | | 0.89 (p<0.001) | 0.01 (p=0.664) |  |
| Heart failure |  | |  | |  |  | 0.048 |
| No | 1.52 (1.35-1.70) | | 1.43 (1.26-1.62) | | 0.92 (p<0.001) | 0.13 (p<0.001) |  |
| Yes | 1.22 (1.04-1.44) | | 1.15 (0.96-1.37) | | 0.78 (p<0.001) | -0.04 (p=0.171) |  |
| Peripheral artery disease | |  | |  |  |  | 0.519 |
| No | 1.47 (1.32-1.63) | | 1.36 (1.21-1.54) | | 0.86 (p<0.001) | 0.11 (p<0.001) |  |
| Yes | 1.24 (1.03-1.49) | | 1.26 (1.03-1.55) | | 0.87 (p<0.001) | -0.03 (p=0.285) |  |

Weights correspond to the inverse probability of having one’s first-line arteriovenous access creation (inverse probability weighting). These probabilities were estimated with logistic regression, including as covariates: age, sex, year of hemodialysis start, primary kidney disease, diabetes, heart failure, peripheral artery disease, coronary heart disease, dysrhythmias, active malignancy, respiratory disease, liver disease, behavior disorders, mobility status, body mass index, urgent and intensive care unit dialysis start, kidney transplant waitlist status, and timing of AV access.

Hazard ratios of hospitalizations were estimated with joint weighted semiparametric frailty models. Each joint model included a submodel for the hazard of recurrent hospitalizations (all cause or cause-specific hospitalizations) and a submodel for the hazard of all cause death, which was considered as informative censoring.

*****Median variance of random effect and alpha in the weighted model through 36 imputed datasets. Positives values of alpha indicate that vascular access-related hospitalization and death are positively correlated; values not significantly different of zero indicate no correlation between these two events.

### Table S5. Hazard ratios (95% confidence intervals) of cardiovascular-related hospitalization associated with first-line AV graft creation (versus AV fistula), overall and by subgroup.

| Sample | Cardiovascular-related hospitalization | | | | Variance of random effect* | | Alpha* | *P* for interaction  (weighted HR) |
| --- | --- | --- | --- | --- | --- | --- | --- | --- |
|  | Unweighted | | Weighted | |  |  |  |  |
| All patients | 1.31 (1.01-1.70) | | 1.16 (1.05-1.29) | | 1.09 (p<0.001) | | 0.43 (p<0.001) | - |
| Age |  | |  | |  | |  | 0.690 |
| < 70 | 1.23 (1.05-1.44) | | 1.15 (0.96-1.37) | | 1.26 (p<0.001) | | 0.39 (p<0.001) |  |
| 70-79 | 1.26 (1.07-1.48) | | 1.26 (1.06-1.51) | | 1.02 (p<0.001) | | 0.39 (p<0.001) |  |
| ≥ 80 | 1.17 (1.01-1.35) | | 1.12 (0.95-1.31) | | 0.87 (p<0.001) | | 0.51 (p<0.001) |  |
| Sex |  | |  | |  | |  | 0.429 |
| Women | 1.26 (1.10-1.45) | | 1.23 (1.06-1.43) | | 1.06 (p<0.001) | | 0.43 (p<0.001) |  |
| Men | 1.22 (1.08-1.37) | | 1.14 (0.99-1.30) | | 1.11 (p<0.001) | | 0.43 (p<0.001) |  |
| Diabetes |  | |  | |  | |  | 0.405 |
| No | 1.31 (1.14-1.49) | | 1.24 (1.07-1.44) | | 1.06 (p<0.001) | | 0.46 (p<0.001) |  |
| Yes | 1.12 (0.99-1.26) | | 1.12 (0.93-1.35) | | 1.00 (p<0.001) | | 0.42 (p<0.001) |  |
| Heart failure |  | |  | |  | |  | 0.352 |
| No | 1.32 (1.18-1.48) | | 1.23 (1.09-1.40) | | 1.12 (p<0.001) | | 0.41 (p<0.001) |  |
| Yes | 1.07 (0.78-1.45) | | 1.07 (0.82-1.40) | | 0.92 (p<0.001) | | 0.39 (p<0.001) |  |
| Peripheral artery disease | |  | |  | |  | | 0.147 |
| No | 1.27 (1.14-1.42) | | 1.19 (1.14-1.24) | | 1.14 (p<0.001) | | 0.44 (p<0.001) |  |
| Yes | 0.99 (0.85-1.15) | | 1.05 (0.89-1.24) | | 0.69 (p<0.001) | | 0.36 (p<0.001) |  |

Weights correspond to the inverse probability of having one’s first-line arteriovenous access creation (inverse probability weighting). These probabilities were estimated with logistic regression, including as covariates: age, sex, year of hemodialysis start, primary kidney disease, diabetes, heart failure, peripheral artery disease, coronary heart disease, dysrhythmias, active malignancy, respiratory disease, liver disease, behavior disorders, mobility status, body mass index, urgent and intensive care unit dialysis start, kidney transplant waitlist status, and timing of AV access.

Hazard ratios of hospitalizations were estimated with joint weighted semiparametric frailty models. Each joint model included a submodel for the hazard of recurrent hospitalizations (all cause or cause-specific hospitalizations) and a submodel for the hazard of all cause death, which was considered as informative censoring.

*****Median variance of random effect and alpha in the weighted model through 36 imputed datasets. Positives values of alpha indicate that cardiovascular-related hospitalization and death are positively correlated.

### Table S6. Hazard ratios (95% confidence intervals) of infection-related associated with first-line AV graft creation (versus AV fistula), overall and by subgroup.

| Sample | Infection-related hospitalization | | | | Variance of random effect* | | Alpha* | *P* for interaction |
| --- | --- | --- | --- | --- | --- | --- | --- | --- |
|  | Unweighted | | Weighted | |  |  |  | (weighted HR) |
| All patients | 1.24 (1.10-1.40) | | 1.16 (1.01-1.33) | | 1.37 (p<0.001) | | 0.44 (p<0.001) | - |
| Age |  | |  | |  | |  | 0.931 |
| < 70 | 1.32 (1.06-1.63) | | 1.17 (0.91-1.50) | | 1.92 (p<0.001) | | 0.47 (p<0.001) |  |
| 70-79 | 1.19 (0.98-1.45) | | 1.18 (0.97-1.44) | | 0.93 (p<0.001) | | 0.66 (p<0.001) |  |
| ≥ 80 | 1.19 (0.98-1.45) | | 1.12 (0.90-1.39) | | 1.06 (p<0.001) | | 0.33 (p<0.001) |  |
| Sex |  | |  | |  | |  | 0.014 |
| Women | 1.39 (1.16-1.67) | | 1.45 (1.17-1.78) | | 1.32 (p<0.001) | | 0.31 (p<0.001) |  |
| Men | 1.16 (0.99-1.35) | | 1.02 (0.86-1.22) | | 1.42 (p<0.001) | | 0.52 (p<0.001) |  |
| Diabetes |  | |  | |  | |  | 0.024 |
| No | 1.41 (1.18-1.67) | | 1.37 (1.12-1.67) | | 1.50 (p<0.001) | | 0.51 (p<0.001) |  |
| Yes | 1.10 (0.93-1.29) | | 0.98 (0.83-1.20) | | 1.24 (p<0.001) | | 0.35 (p<0.001) |  |
| Heart failure |  | |  | |  | |  | 0.291 |
| No | 1.36 (1.17-1.58) | | 1.26 (1.06-1.49) | | 1.52 (p<0.001) | | 0.49 (p<0.001) |  |
| Yes | 1.03 (0.84-1.27) | | 1.01 (0.74-1.40) | | 1.01 (p<0.001) | | 0.32 (p<0.001) |  |
| Peripheral artery disease | |  | |  | |  | | 0.309 |
| No | 1.29 (1.12-1.50) | | 1.21 (1.02-1.42) | | 1.49 (p<0.001) | | 0.43 (p<0.001) |  |
| Yes | 1.07 (0.86-1.33) | | 1.04 (0.82-1.31) | | 0.96 (p<0.001) | | 0.44 (p<0.001) |  |

Weights correspond to the inverse probability of having one’s first-line arteriovenous access creation (inverse probability weighting). These probabilities were estimated with logistic regression, including as covariates: age, sex, year of hemodialysis start, primary kidney disease, diabetes, heart failure, peripheral artery disease, coronary heart disease, dysrhythmias, active malignancy, respiratory disease, liver disease, behavior disorders, mobility status, body mass index, urgent and intensive care unit dialysis start, kidney transplant waitlist status, and timing of AV access.

Hazard ratios of hospitalizations were estimated with joint weighted semiparametric frailty models. Each joint model included a submodel for the hazard of recurrent hospitalizations (all cause or cause-specific hospitalizations) and a submodel for the hazard of all cause death, which was considered as informative censoring.

*****Median variance of random effect and alpha in the weighted model through 36 imputed datasets. Positives values of alpha indicate that infectious-related hospitalization and death are positively correlated.

### Table S7. Hazard ratios (95% confidence intervals) of all-cause death associated with first-line AV graft creation (versus AV fistula), overall and by subgroup.

| Sample | All-cause death | |  |
| --- | --- | --- | --- |
|  | Unweighted | Weighted | *P* for interaction (weighted HR) |
| All patients | 1.33 (1.20-1.47) | 1.09 (0.97-1.23) | - |
| Age |  |  | 0.210 |
| < 70 | 1.27 (1.03-1.57) | 1.07 (0.84-1.37) |  |
| 70-79 | 1.41 (1.18-1.67) | 1.24 (1.03-1.51) |  |
| ≥ 80 | 1.08 (0.93-1.26) | 0.99 (0.83-1.17) |  |
| Sex |  |  | 0.512 |
| Women | 1.29 (1.10-1.52) | 1.03 (0.86-1.24) |  |
| Men | 1.37 (1.21-1.56) | 1.12 (0.96-1.30) |  |
| Diabetes |  |  | 0.369 |
| No | 1.42 (1.23-1.65) | 1.15 (0.97-1.36) |  |
| Yes | 1.23 (1.07-1.41) | 1.03 (0.87-1.21) |  |
| Heart failure |  |  | 0.205 |
| No | 1.26 (1.11-1.45) | 1.04 (0.89-1.22) |  |
| Yes | 1.31 (1.13-1.54) | 1.21 (1.02-1.45) |  |
| Peripheral artery disease |  | | 0.035 |
| No | 1.41 (1.25-1.59) | 1.16 (1.00-1.34) |  |
| Yes | 1.03 (0.86-1.25) | 0.88 (0.70-1.09) |  |

Weights correspond to the inverse probability of having one’s first-line arteriovenous access creation (inverse probability weighting). These probabilities were estimated with logistic regression, including as covariates: age, sex, year of hemodialysis start, primary kidney disease, diabetes, heart failure, peripheral artery disease, coronary heart disease, dysrhythmias, active malignancy, respiratory disease, liver disease, behavior disorders, mobility status, body mass index, urgent and intensive care unit dialysis start, kidney transplant waitlist status, and timing of AV access.

Hazard ratios of death, regardless of whether death was preceded by any hospitalization, were estimated with Cox proportional hazard models.

### Table S8. Sensitivity analysis: Assessment of the balance in the frequency of preoperative imaging and the number of hospitalizations in the two years preceding arteriovenous access creation, by diabetes status.

|  | AV fistula | AV graft | Absolute standardized difference | | |
| --- | --- | --- | --- | --- | --- |
|  |  |  | Unweighted | Weighted  (main analysis PS) | Weighted  (sensitivity analysis PS) |
| **Without diabetes** |  |  |  |  |  |
| Preoperative imaging before AV access creation, % | 32 | 46 | 0.14 | 0.14 | 0.03 |
| Number of hospitalizations in the two years preceding AV access creation, mean (SD) | 3.9 (2.7) | 4.5 (3.1) | 0.23 | 0.19 | 0.03 |
| **With diabetes** |  |  |  |  |  |
| Preoperative imaging before AV access creation, % | 36 | 46 | 0.10 | 0.09 | 0.03 |
| Number of hospitalizations in the two years preceding AV access creation, mean (SD) | 4.6 (3.2) | 5.4 (3.5) | 0.23 | 0.15 | 0.01 |

Weights correspond to the inverse probability of having one’s first-line arteriovenous access creation (inverse probability weighting). These probabilities were estimated with logistic regression, including as covariates: age, sex, year of hemodialysis start, primary kidney disease, diabetes, heart failure, peripheral artery disease, coronary heart disease, dysrhythmias, active malignancy, respiratory disease, liver disease, behavior disorders, mobility status, body mass index, urgent and intensive care unit dialysis start, kidney transplant waitlist status, and timing of AV access.

Abbreviations: AV, arteriovenous; PS, propensity score; SD, standard deviation.

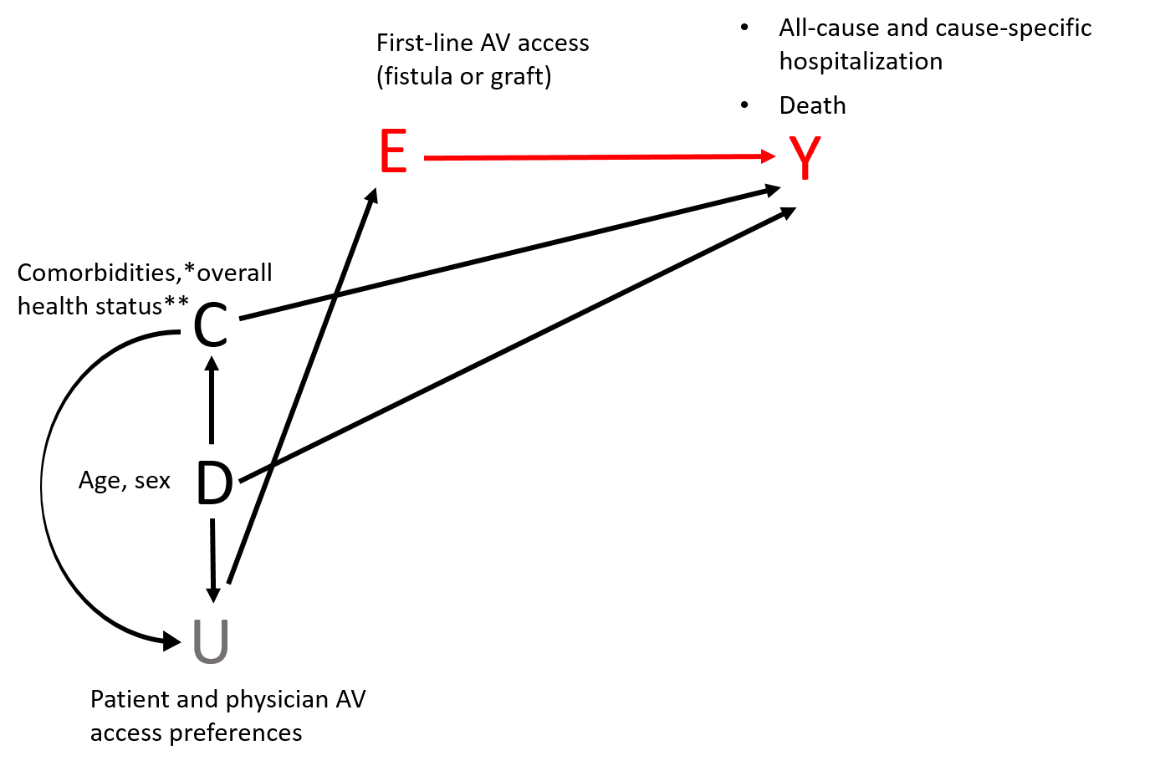

**Figure S1. Direct acyclic graph for the relation between first-line arteriovenous access and outcomes.**

In this direct acyclic graph, age, sex (“D”), comorbidities, and overall health status reflected in the REIN registry by a number of indicators specified below (“C”) are considered to affect the first-line AV access type (“E”) through the unobserved patient and physician AV access preferences (“U”). “D”, “C”, and “E” are also considered to affect hospitalization and death hazards (“Y”). We hypothesize that patient and physician AV access preferences do not affect patient outcomes otherwise then by the first-line AV access created (intention-to-treat approach), so that adjusting for “C” and “D” is enough to obtain a causal effect of the first-line AV access on subsequent hospitalization and death.

*Primary kidney disease, diabetes, heart failure stages according to the New York Heart Association (NYHA) Classification, peripheral artery disease stages according to the Leriche-Fontaine Classification, coronary heart disease, dysrhythmias, active malignancy, respiratory disease, liver disease, behavioral disorders.

**Reflected in the REIN Registry by the time from dialysis start to arteriovenous access creation, body mass index, mobility status, urgency dialysis start, dialysis start in an intensive care unit, and inscription in the kidney transplant waitlist.

Abbreviations: AV, arteriovenous; Tx, transplant.

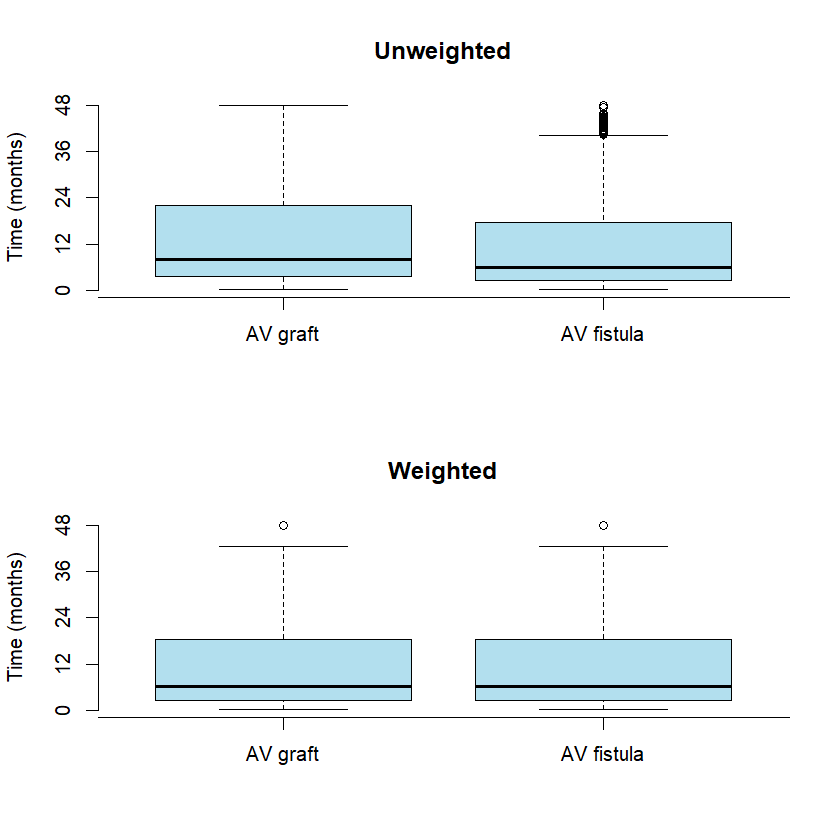

**Figure S2. Unweighted and weighted distributions of time** **from dialysis start to arteriovenous access creation.**

Weights correspond to the inverse probability of having one’s first-line arteriovenous access creation (inverse probability weighting). These probabilities were estimated with logistic regression, including as covariates: age, sex, year of hemodialysis start, primary kidney disease, diabetes, heart failure, peripheral artery disease, coronary heart disease, dysrhythmias, active malignancy, respiratory disease, liver disease, behavior disorders, mobility status, body mass index, urgent and intensive care unit dialysis start, kidney transplant waitlist status, and timing of AV access.

Abbreviation: AV, arteriovenous.

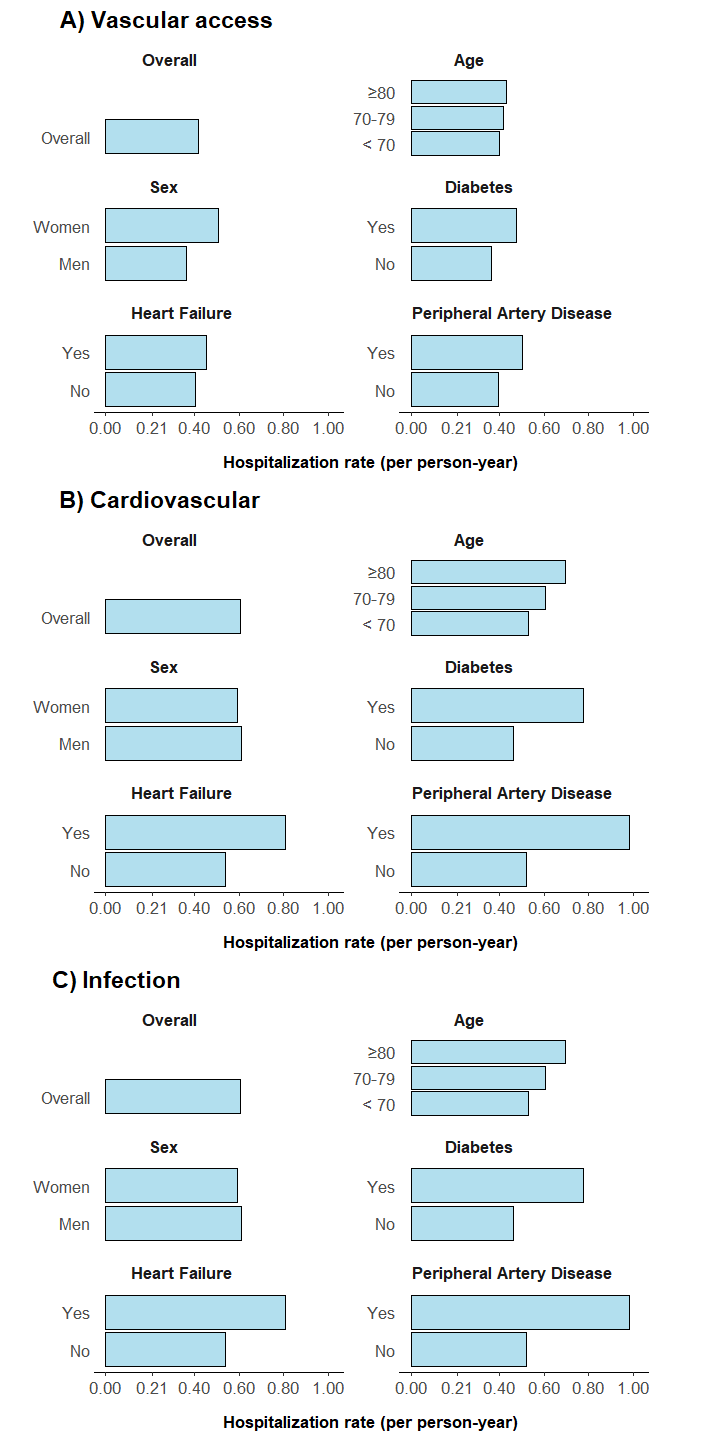

**Figure S3. Vascular access- (A), cardiovascular- (B), and infection- related (C) hospitalization rates in the overall study population and across subgroups of age, sex, diabetes, heart failure, and peripheral artery disease.**

Weights correspond to the inverse probability of having one’s first-line arteriovenous access creation (inverse probability weighting). These probabilities were estimated with logistic regression, including as covariates: age, sex, year of hemodialysis start, primary kidney disease, diabetes, heart failure, peripheral artery disease, coronary heart disease, dysrhythmias, active malignancy, respiratory disease, liver disease, behavior disorders, mobility status, body mass index, urgent and intensive care unit dialysis start, kidney transplant waitlist status, and timing of AV access.
